# Environmental predominance on brain structure and connectivity in language-related regions: a multimodal twin study in adolescence

**DOI:** 10.64898/2026.09.30.26364387

**Authors:** Emma Tassi, Chiara Colli, Aida Peyvandi, Lorenza De Nardi, Camilla Pansitta, Silvia Busilacchi, Francesca Marzia Siri, Giuseppe Delvecchio, Eleonora Maggioni, Carolina Bonivento, Maddalena Mauri, Corrado Fagnani, Emanuela Medda, Maria Nobile, Paolo Brambilla

**Affiliations:** Department of Neurosciences and Mental Health, Fondazione IRCCS Ca’ Granda Ospedale Maggiore Policlinico, Milan, Italy; Department of Pathophysiology and Transplantation, Università degli Studi di Milano, Milan, Italy; Department of Electronics, Information and Bioengineering, Politecnico di Milano, Milan, Italy; Polo FVG, Scientific Institute IRCCS ‘Eugenio Medea’, Pasian di Prato, Udine, Italy; Child Psychopathology Unit, Scientific Institute IRCCS Eugenio Medea, Bosisio Parini, Italy; Centre for Behavioural Sciences and Mental Health, Istituto Superiore di Sanità, Rome, Italy

**Keywords:** Twin studies, Multimodal neuroimaging, Language-related networks, Psychopathological risk, Heritability, Environmental factors

## Abstract

Language alterations are recognized as early markers of psychopathological vulnerability, yet genetic and environmental factors underlying language-related brain regions remain unknown. We applied univariate twin analyses and risk-discordant comparisons to multimodal dataset from 31 healthy twin pairs (9–31 years) and 17 pairs discordant for psychopathological risk (10–27 years). Gray matter volume (GMV), cortical thickness (CT), and functional interactions, specifically effective connectivity (EC), were assessed in eight bilateral language-related regions, alongside linguistic, emotional-behavioural, temperamental, and personality-related phenotypes. In healthy twin pairs (27–31 pairs, depending on data completeness), genetic and environmental determinants were estimated for all phenotypes. In 17 pairs discordant for psychopathological risk, intra-pair neuroimaging differences were tested for association with psychopathological discordance and zygosity, and individual brain measures were compared across high-risk twins, their low-risk co-twins, and healthy controls. Environmental factors predominated in structural and functional brain variability, while genetic contributions were limited, region-specific. Narrative fluency and psychological traits showed stronger heritability. Findings converged on the right middle frontal gyrus, where GMV and CT alterations reflected non-shared environment and risk expression. Despite the small sample, preliminary results suggest neurobiological vulnerability in language networks arises from environmental rather than genetic factors, informing environmental susceptibility to adolescent mental illness.

## 1. Introduction

Language is a fundamental and complex human ability that emerges from the dynamic interaction of several brain areas. It plays a pivotal role in social connections and work functioning[1], and from a clinical standpoint it represents one of the primary tools for evaluating patients’ mental states.

Thinking disorders are primarily observable through linguistic disruptions [2,3], which have long been regarded as hallmark symptoms of psychosis [4–6]. However, accumulating evidence suggests that linguistic alterations are not confined to specific diagnostic categories but occur across a broad range of psychiatric conditions, encompassing both states of vulnerability and fully developed disorders [6–9].

Neuroimaging studies have begun to elucidate the neural substrates underlying these disturbances, pointing to specific dysfunctions within language-related brain networks [10]. The most prominent alterations have been reported in the inferior frontal gyrus (IFG), the superior temporal gyrus (STG), and the prefrontal cortex (PFC) [11,12]. These findings are consistent with evidence from a large meta-analysis of 403 neuroimaging experiments in healthy adults, showing that language processing primarily engages bilateral fronto-temporal cortices, with consistent activation in the left IFG, the right anterior IFG extending into the insula, the left superior and middle temporal gyri, and the middle portions of the right STG and posterior middle temporal gyrus (MTG) [13].

Despite this growing understanding of the neural architecture of language, the extent to which language-related brain features are shaped by genetic and environmental influences remains largely unexplored. Twin studies provide an excellent platform for estimating these contributions, by comparing intra-pair correlations between monozygotic (MZ) and dizygotic (DZ) twins and to decompose phenotypic variance into additive genetic (A), shared environmental (C), unique environmental (E), and nonadditive genetic (D) components through ACE and ADE phenotypic decomposition of variance [14–18].

Available neuroimaging evidence suggests that genetic and environmental influences vary substantially across imaging phenotypes, brain regions, and developmental stages. Cortical thickness (CT) exhibits region-specific heritability rather than a uniform genetic architecture [19], and substantial heterogeneity in genetic expression has been documented across cortical regions and developmental periods [20]. Twin studies further indicate that CT and white matter connectivity may share genetic influences within selected networks [21], whereas frontal and temporal cortical regions often appear particularly sensitive to environmental modulation [22]. Longitudinal twin investigations have additionally shown that developmental changes in cortical and subcortical structures are shaped by both genetic and environmental factors throughout childhood and adolescence [23,24]. In this context, our previous twin study focused on resting-state functional connectivity, exploring genetic and environmental influences across a wide range of functional network features at multiple spatial scales, and showed a predominant contribution of unique environmental factors to inter-individual variability in brain functional connectivity [18].

At the linguistic level, twin studies suggest that genetic and environmental contributions to language are dynamic across development. Oral language abilities share substantial genetic overlap with reading comprehension, whereas productive language appears to be predominantly influenced by non-shared environmental factors [25,26]. Furthermore, language-related skills have been shown to exhibit moderate heritability across childhood and adolescence [27], supporting a genetic contribution to higher-order language abilities [28].

Similarly, emotional-behavioural traits arise from varying combinations of genetic and environmental influences. Twin studies based on the Child Behavior Checklist (CBCL) have consistently reported moderate-to-high heritability for internalizing, externalizing, and DSM-oriented symptom dimensions, while also highlighting substantial contributions of shared environmental factors for selected behavioural domains [29–31]. Longitudinal evidence further indicates that genetic influences contribute substantially to the stability of internalizing and externalizing behaviours across development, whereas environmental exposures may shape their trajectories over time [32]. However, no study has simultaneously investigated the genetic and environmental determinants of multimodal brain features and linguistic phenotypes within language-related regions during adolescence and young adulthood.

Understanding how language alterations relate to anatomical and functional connectivity in brain morphology and functional connectivity within language-related networks during the early stages of psychopathology is particularly relevant, as it may help identify clinical and brain markers that precede illness onset. In this context, the discordant twin design represents a particularly informative approach. By comparing twin pairs in which only one twin carries the target condition, this design can help disentangle etiological factors and characterize intermediate phenotypes.

Twin MRI studies in schizophrenia have demonstrated that some gray matter abnormalities track illness or unique environmental exposure beyond genetic liability alone [33,34], highlighting the potential of discordant-twin designs for understanding genetic and environmental influences on brain vulnerability. Moreover, the inclusion of an external control group matched to the discordant twins for age and sex can provide additional insight into the relative weight of genetic and environmental factors acting on a given trait of interest [35].

This study aimed to clarify how genetic and environmental factors shape language-related brain regions during adolescence and young adulthood, a critical developmental window for the emergence of psychopathological vulnerability, and whether these brain features are associated with psychopathological risk.

Specifically, this study aimed to: (i) quantify genetic and environmental influences on psychopathological risk-related traits, linguistic features, and multimodal measures extracted from language-related brain regions in healthy twins; and (ii) identify language-related brain phenotypes associated with psychopathological risk through the analysis of risk-discordant twin pairs.

To this aim, we conducted a multimodal and multilevel twin analysis to disentangle genetic and environmental influences on psychological, linguistic, and brain features during adolescence and young adulthood. Brain measures included regional morphology —gray matter volume (GMV) and CT— and causal interactions —captured by effective connectivity (EC)— examined across eight bilateral language-related regions of interest. The selection of these ROIs was guided by a comprehensive literature review targeting brain regions most robustly associated with language processing across linguistic subdomains. Key areas included the IFG, STG, and MFG [4,13]. Within the inferior frontal lobe, the pars opercularis and pars triangularis were included as critical hubs for semantic and syntactic processing [4,36].

## 2. Methods

### 2.1 Participants and procedure

The present work is part of the LearningTwin project (Italian MoH, SG-2021-12375585), which is ongoing at Fondazione IRCCS Cà Granda Ospedale Policlinico of Milan. The dataset analysed was previously collected from a young twin sample at the scientific Institute IRCCS E. Medea (SPES study, Italian MoH, GR-2010-2316745; WhyMe? Study, Italian MoH, RF-2011-02352308), within the same research framework and recruitment protocol described in Tassi et al., 2023[18]. The cohort included healthy control twin pairs (HC) and twin pairs discordant for psychopathological risk (risk-discordant), recruited through the Italian Twin Registry[37]. Exclusion criteria were intelligence quotient (IQ) < 70 based on Wechsler intelligence scales for adults/children (WAIS/WISC), diagnosis of autism spectrum condition, epilepsy or other neurological disorders, history of head trauma, severe visual, auditory, or language comprehension problems, and gestational age < 34 weeks. The original data collection protocol (SPES and WhyMe? studies) was approved by the Ethics Committee of IRCCS “Eugenio Medea” - Associazione “La Nostra Famiglia”, in accordance with the 2013 Fortaleza version of the Declaration of Helsinki and subsequent amendments. The retrospective use of these data within the present LearningTwin project (SG-2021-12375585) was approved by the Comitato Etico Milano Area 2, of Fondazione IRCCS Cà Granda Ospedale Maggiore Policlinico. Written informed consent to the study was obtained from all participants; for minor twin participants under 18 years of age, consent was provided by their parents and/or legal guardian.

The sample selected for this study comprised same-sex HC and risk-discordant twin pairs, including MZ and DZ twins. Risk-discordant twin pairs were defined as those pairs in which only one twin met criteria for psychotic and/or emotional-behavioural risk based on standardized assessments. Specifically, subjects were considered at risk if at least one of the three dimensions of Child/Adult Behaviour Checklist (CBCL/ABCL) scale was above the cutoff of T-score=60 (internalizing, externalizing, or total subscales > 60), indicating emotional-behavioural alterations, or if early psychotic risk was positive based on the ERIraos Checklist (Pre-psychosis: YES) [38,39].

### 2.2 Measurements

The original dataset comprised sociodemographic, psychopathological, behavioural, cognitive, and multimodal brain magnetic resonance imaging (MRI) data. In the current study, psychological, linguistic and multimodal neuroimaging features were considered. Specifically, the analysis considered multimodal features including 17 psychological features spanning emotional-behavioural symptoms, temperament, personality traits, and subclinical psychotic traits; 8 linguistic features describing narrative language abilities at both micro- and macro-linguistic levels; 16 brain morphological features; and 8 brain functional features.

#### 2.2.1 Psychological measures

We considered in total seventeen psychological scale scores derived from the following measures: (i) ABCL/CBCL scales including (a) seven syndrome scales empirically derived from factorial analyses, including Anxious/Depressed, Withdrawn/Depressed, Somatic Complaints, Social Problems (only for CBCL), Thought Problems, Attention Problems, Rule-Breaking Behaviour (only for ABCL), (b) three second-order scales summarizing Internalizing Problems (Anxiety/ Depressed, Withdrawn/Depressed, and Somatic Complaints), Externalizing Problems (Aggressive and rule-breaking Behaviours), and Total Problems; (ii) Temperament and Character Inventory (TCI) scale including four dimensions of temperament (Novelty Seeking, Harm Avoidance, Reward Dependence, and Persistence); (iii) Personality Assessment Scale (PAS); (iv) Magical Ideation Scale (MIS); (v) Hypomanic Personality Scale (HPS), (vi) ERIraos Checklist (ERIraos-CL). Specifically, CBCL/ABCL and ERIraos represent the psychological measures used to define the psychopathological risk discordance (i.e., psychotic and/or emotional-behavioural risk) between twins in the risk discordant group. Detailed descriptions of each psychological scale, including psychometric properties and normative references, are reported in Supplementary Materials (Methods section).

#### 2.2.2 Linguistic features

Based on the study of Gargano et al. (2022) and Marini et al. (2011), nine linguistic features were considered in this study, including words, narrative fluency, phonological paraphasia, semantic paraphasia, lexical informativeness, paragrammatical errors, syntactic completeness, local coherence errors, and global coherence errors [4,40]. These features were used to measure participants’ narrative language skills. Specifically, they measure micro-linguistic elements (i.e., words, narrative fluency, phonological and semantic paraphasias, paragrammatical errors, syntactic completeness), and macro-linguistic structures (i.e., lexical informativeness, syntactic completeness, local coherence errors, and global coherence errors).

#### 2.2.3 Neuroimaging features

##### MRI acquisition protocol

Structural and resting-state functional MRI data were acquired on a 3T Philips Achieva scanner (Philips, Best, The Netherlands) with a multi-channel head coil at IRCCS E. Medea, using the same acquisition protocol adopted for this twin cohort in earlier work[18]. Structural images (sMRI) were collected with a 3D T1-weighted turbo field echo (TFE) sequence combined with SENSE parallel imaging (repetition time (TR) = 8.1 ms, echo time (TE) = 3.7 ms, flip angle = 8°, 190 contiguous axial slices, in-plane matrix = 240 × 240, isotropic voxel size = 1 × 1 × 1 mm³, SENSE factor = 2). For resting-state functional MRI (rs-fMRI, 200 volumes), a T2*-weighted gradient-echo echo-planar sequence was used (TR = 2500 ms, TE = 35 ms, flip angle = 8.5°, 30 contiguous axial slices, in-plane matrix = 128 × 128, slice thickness = 3 mm, SENSE factor = 2). During the rs-fMRI scan, participants were instructed to keep their eyes closed, remain still, avoid deliberate thought, and stay awake.

##### sMRI processing and feature extraction

T1-weighted sMRI images were processed using the Computational Anatomy Toolbox (CAT-12: https://neuro-jena.github.io/cat/mi)[41], an advanced extension of the Statistical Parametric Mapping software (SPM12: http://www.fil.ion.ucl.ac.uk/spm/)[42], to derive regional GMV and CT. Processing was performed as previously described [43], and briefly comprised: segmentation of brain tissue classes, spatial normalization to the MNI template, volumetric correction of the segmented tissue for registration-related distortions (modulation), and spatial smoothing with a 3D Gaussian kernel (6 mm FWHM). The resulting smoothed, modulated, normalized GM maps were then used to derive GMV and CT values across regions defined by probabilistic atlases. For each subject, mean GMV values were computed for the volume-based Neuromorphometrics atlas (n=136) [44], and mean CT for the surface-based Desikan-Killiany atlas (n=72) [45]. From these parcellations we retained only the regions belonging to the language network of interest, including the middle frontal gyrus (MFG), the pars opercularis and pars triangularis of the inferior frontal gyrus (IFG), and the superior temporal gyrus (STG), represented bilaterally. This selection was guided by previous literature identifying fronto-temporal regions as core components of language processing, with the pars opercularis and pars triangularis included as key regions for semantic and syntactic processing [4,13,36].

##### fMRI preprocessing and feature extraction

The fMRI pre-processing was performed using HALfpipe[46], a standardized ENIGMA-consortium workflow for task-based and resting-state fMRI, following a pipeline coherent with our previous work [47]. Briefly, this comprised realignment of functional volumes, co-registration to the corresponding T1-weighted image, tissue segmentation, and normalization to MNI space. Standard additional steps were also applied, namely spatial smoothing (6 mm FWHM), grand-mean intensity scaling (mean = 10,000), and Gaussian-weighted temporal filtering (0–125 s band). Denoising further included regression of motion parameters and physiological signals, individually and in combination [48]. Head motion was quantified via framewise displacement (FD), which averaged 0.12 mm (± 0.07 mm) across participants, under the standard 0.5 mm[49]. Regional time series were then extracted with the SPM Marsbar toolbox (version 0.44, http://marsbar.sourceforge.net/)[50], parcellating rs-fMRI volumes according to the Automated Anatomical Labeling (AAL) atlas and restricting extraction to the same language-network ROIs used for the structural analyses (bilateral MFG, pars opercularis and pars triangularis of the IFG, and STG), allowing structural and functional features to be analysed within a shared language-related areas[4,13,36].

Effective connectivity (EC) among the selected regions was estimated via Granger Causality Analysis (GCA), adapting functions from the MATLAB Granger Multivariate Autoregressive Connectivity (GMAC) toolbox[51], following an approach consistent with previous work[52]. For each participant, a multivariate autoregressive (MVAR) model of order 2, the average of the individually optimal orders obtained via the Akaike Information Criterion (AIC), was fitted to the fMRI time series of each selected ROI, treated as a network node. Partial Directed Coherence (PDC) [53] was then computed from the resulting MVAR coefficients⁵³, providing a frequency-domain estimate of the strength and direction of inter-regional information transfer. Given a region pair (i, j), PDC captures the directed influence exerted by region i on region j, and viceversa. Connection-level significance was tested against a surrogate-based null distribution, generated by recomputing PDC on 200 randomized datasets; observed values were compared to this distribution with one-tailed tests, applying a Bonferroni correction (p < 0.05) across the 56 directed regional pairs considered. Connections that did not reach significance were set to zero, producing the final 8-node EC network. From this network, we derived three summary measures per node, capturing its role in the directional flow of information: outflow (out-degree; number of significant outgoing connections), inflow (in-degree; number of significant incoming connections), and NetFlow (outflow minus inflow), reflecting whether a region acted predominantly as a source or a target of information. All subsequent analyses focused on NetFlow, as the single measure best summarizing each region’s overall directional role within the language-related network.

The EC network of the 8 selected ROIs was constructed by retaining significant PDC values and setting non-significant connections to zero. From the resulting network, we derived node-level summary measures by aggregating the strength of incoming and outgoing information flow: Outflow (out-degree), the number of significant causal connections from a node to other nodes; Inflow (in-degree), the number of significant causal connections to a node from other nodes; and NetFlow (out–in degree), the difference between outflow and inflow, capturing the net directional influence of each node within the network. For all subsequent analyses, we focused exclusively on the NetFlow parameter, as it provides a single summary measure of each region’s net causal role – whether predominantly sending or receiving information – within the language-related network.

### 2.3 Genetic and Environmental Influences on features

Statistical analyses were performed using MATLAB R2025a. The following steps were performed.

#### 2.3.1 Intra-pair feature correlation

For each feature (i.e., psychological, multimodal neuroimaging, linguistic), separate analyses were conducted for MZ and DZ twin pairs. The partial correlation coefficient between the feature values of Twin #1 and Twin #2 was computed, controlling for age and sex. As a result, we obtained zygosity-specific correlations, rMZ and rDZ. These correlations were compared to determine the most appropriate genetic model to fit, as described in the following section.

For all features, regardless of the most suitable genetic-environmental variance decomposition, significant differences between rMZ and rDZ were assessed based on their Fisher’s Z-transformation, with significance threshold set at p = 0.05. The resulting Z-statistics and associated p-values are reported in **Tables 2-3**.

**Table 1.** Demographic and behavioural information of the primary HC subsample (N=27 twin pairs with complete multimodal data) and risk-discordant sample (N=17 twin pairs)

| Variable | HC MZ<br>(11 pairs) | HC DZ<br>(16 pairs) | HC Stats | Risk-discordant<br>MZ<br>(9 pairs) | Risk-discordant<br>DZ<br>(8 pairs) | Risk-discordant Stats |
| --- | --- | --- | --- | --- | --- | --- |
| **Twin pairs** | 11 | 16 | — | 9 | 8 | — |
| **Age (years)** | 14.59 ± 4.66 | 15.18 ± 4.58 | $T(z) = -0.22$, $p = 0.80$<br>(pair-based) | 15.14 ± 5.93 | 14.82 ± 5.06 | $T(z) = -0.28$, $p = 0.81$<br>(pair-based) |
| **Sex pairs**<br>**(M/F)** | 8/3 | 4/12 | $\chi^2 = 6.04$, $p = 0.002$ | 7/2 | 3/5 | $\chi^2 = 2.83$, $p = 0.09$ |
| **ERlraos-CL** | 2.09 ± 2.95 | 2.19 ± 1.91 | — | 5.00 ± 8.92 | 6.12 ± 8.72 | T(z) = -0.67, p = 0.52 (pair-based) |
| **ABCL/CBCL Total** | 45.55 ± 5.13 | 46.50 ± 3.90 | T(z) = -0.54, p = 0.60 (pair-based) | 50.75 ± 5.09 | 59.00 ± 3.51 | T(z) = -2.80, p = 0.002 (pair-based) |

Abbreviations: HC, healthy controls; ABCL: Adult Behavior Checklist; CBCL: Child Behavior Checklist; DZ: Dizygotic twins; ERlraos: Early Recognition Inventory for the Retrospective Assessment of the Onset of Schizophrenia; MZ: Monozygotic twins. T: T-statistics; $\chi^2$ : $\chi^2$ statistics; p: p-value.

**Table 2.** Statistical twin analyses results for Psychological and linguistic features in healthy twin pairs (N = 29 twin pairs for psychological; N = 31 twin pairs for linguistic).

| Metric domain | Variable | rMZ | rDZ | Fisher Z-statistics (rMZ vs rDZ) | p-values | Genetic – Environmental parameters |
| --- | --- | --- | --- | --- | --- | --- |
| Linguistic | Narrative Fluency | 0.82 | 0.20 | $Z = 2.27$ | $p = 0.01$ | Epistatic effect; A/C/D/E not estimated |
| Psychological | ABCL/CBCL Total | 0.76 | 0.46 | $Z = 1.17$ | n.s. | A: 0.59, C: 0.17, E: 0.24 |
| | ABCL/CBCL Thought Problems | 0.82 | 0.74 | $Z = 0.46$ | n.s. | A: 0.15, C: 0.66, E: 0.18 |
| | ABCL/CBCL Rule-Breaking | 0.71 | 0.47 | $Z = 0.92$ | n.s. | A: 0.49, C: 0.22, E: 0.29 |
| | MIS | 0.64 | 0.42 | $Z = 0.73$ | n.s. | A: 0.44, C: 0.20, E: 0.36 |
| | HPS | 0.74 | 0.69 | $Z = 0.23$ | n.s. | A: 0.09, C: 0.64, E: 0.26 |
| | ABCL/CBCL Internalizing | 0.52 | 0.19 | $Z = 0.91$ | n.s. | A: 0.23, D: 0.29, E: 0.48 |
| | ABCL/CBCL Somatic Complaints | 0.62 | 0.22 | $Z = 1.20$ | n.s. | A: 0.26, D: 0.36, E: 0.38 |

Abbreviations: rMZ: intraclass correlation in monozygotic twin pairs; rDZ: intraclass correlation in dizygotic twin pairs. A, additive genetic effect; D, dominant genetic effect; C, shared environmental effect; E, unique environmental effect; ABCL: Adult Behavior Checklist; CBCL: Child Behavior Checklist; MIS: Magical Ideation Scale; HPS: Hypomanic Personality Scale; n.s.: not significant For features showing an epistatic pattern, ACE/ADE decomposition was not applicable. Fisher's Z and corresponding p-values refer to the statistical comparison between rMZ and rDZ correlations.

**Table 3.** Statistical twin analyses results for multimodal neuroimaging features in healthy twin pairs (sMRI: N = 29 twin pairs; fMRI: N = 27 twin pairs).

| Metric domain | ROIs | rMZ | rDZ | Fisher Z-statistics (rMZ vs rDZ) | p-values | Genetic – Environmental parameters |
| --- | --- | --- | --- | --- | --- | --- |
| **GMV** | Right MFG | 0.50 | 0.45 | Z=0.16 | n.s. | A: 0.10<br>C: 0.40<br>E: 0.50 |
|  | Left Opercular part of IFG | 0.68 | 0.09 | Z=1.74 | p =0.04 | Epistatic effect; A/C/D/E not estimated |
|  | Right Triangular part of the IFG | 0.70 | 0.60 | Z=0.39 | n.s. | A: 0.19<br>C: 0.51<br>E: 0.30 |
|  | Right STG | 0.80 | 0.68 | Z=0.61 | n.s. | A: 0.23<br>C: 0.57<br>E: 0.20 |
|  | Left STG | 0.74 | 0.50 | Z=0.94 | n.s. | A: 0.47<br>C: 0.27<br>E: 0.26 |
| **CT** | Left Opercular part of IFG | 0.61 | 0.17 | Z=1.28 | n.s. | A: 0.08<br>D: 0.53<br>E: 0.39 |
|  | Left STG | 0.77 | 0.61 | Z=0.72 | n.s. | A: 0.31<br>C: 0.46<br>E: 0.23 |
| **Netflow** | Right Triangular part of the IFG | 0.35 | 0.18 | Z=0.42 | n.s. | A: 0.35<br>C: 0.01<br>E: 0.65 |

Abbreviations: ROIs, regions of interest; GMV: Gray Matter Volume; CT: Cortical Thickness; Netflow, functional features of Netflow extracted from Effective Connectivity analysis; MFG: Middle Frontal Gyrus; IFG: Inferior Frontal Gyrus; STG: Superior Temporal Gyrus; n.s.: not significant; rMZ: intraclass correlation in monozygotic twin pairs; rDZ: intraclass correlation in dizygotic twin pairs. A, additive genetic effect; D, dominant genetic effect; C, shared environmental effect; E, unique environmental effect. For regions showing an epistatic pattern, ACE/ADE decomposition was not applicable. Fisher's Z and corresponding p-values refer to the statistical comparison between rMZ and rDZ correlations.

#### 2.3.2 Genetic-environmental influences on each feature

Based on the intraclass correlations estimated in MZ and DZ twins, the Falconer’s formula was applied to decompose the observed phenotypic variance for features satisfying both of the following conditions[18]: (1) rMZ>0 and rDZ>0, and (2) rMZ>rDZ. The observed phenotypic variance was decomposed in: (i) additive genetic factors (A), which reflect the cumulative influence of alleles affecting the phenotype; either (ii) non-additive genetic factors (D), representing interactions between alleles at the same or different loci (dominance and epistasis, respectively), or (iii) shared environmental factors (C), capturing environmental influences common to family members (e.g., conditions shared during early development); and (iv) unique environmental factors (E), encompassing individual-specific exposures such as lifestyle, infections, or traumatic experiences. A decomposition of phenotypic variance including A, C, and E components (ACE decomposition) was applied when rMZ > rDZ, whereas an ADE decomposition was used when the MZ correlation was approximately twice that of DZ twins (rMZ ≈ 2rDZ). When the condition rMZ > 4rDZ > 0 was met, the feature under examination was interpreted as being affected by non-additive (epistatic) genetic influences. Such effects cannot be disentangled using classical twin analyses, as they are not estimable as a linear function of the observed rMZ and rDZ correlations.

### 2.4 Association of multimodal language-related brain phenotypes with psychopathological risk and zygosity

#### 2.4.1 Within-pair neuroimaging variability: association with psychopathological risk and zygosity

The first analysis examined within-pair variability in multimodal neuroimaging features (CT, GMV, and EC features) by assessing how intra-pair differences relate to phenotypic discordance and zygosity. For each ROI, we computed the absolute intra-pair difference in the neuroimaging measure, reflecting how much the two twins within each pair differ in that specific brain feature.

We then applied linear partial correlations to isolate non-shared environmental and genetic influences on within-pair neuroimaging variability

First, we tested the association between intra-pair differences in the neuroimaging measures and psychopathological risk discordance, while controlling for zygosity (MZ vs. DZ). A significant positive association indicates that risk-discordant pairs show greater neuroimaging dissimilarity than concordant HC pairs, consistent with the influence of non-shared environmental or individual-specific factors, as this pattern emerges when genetic similarity is held constant. Second, we tested the association between intra-pair differences in neuroimaging measures and zygosity, while controlling for psychopathological risk discordance. A significant association in this test suggests that the degree of genetic sharing influences how similar co-twins are in that feature, independently of their discordance status, consistent with genetic contributions to neuroimaging variability.

Significant results were additionally visually inspected using boxplots, displaying the distribution of intra-pair differences across HC and risk-discordant pairs.

#### 2.4.2 Across-group neuroimaging comparison among high-risk, low-risk and healthy individuals

The second analysis aimed to disentangle genetic and environmental contributions to psychopathological risk by comparing all multimodal neuroimaging measures (GMV, CT, and node-level EC) across HC, LR, and HR twins, characterizing whether each feature reflects a familial vulnerability or an individual-specific factor differentiating co-twins.

For each brain modality and each ROI, a Wilcoxon signed-rank test was conducted between paired data of HR and LR individuals (HR vs LR) as they belong to the same twin pairs and a Mann-Whitney test was used to compare unpaired data of HR and LR individuals separately with the HC group (HR vs HC, LR vs HC). Group differences were interpreted according to four theoretically informed patterns that reflect distinct sources of influence.

A significant difference between HR and LR twins, with neither group showing significant differences from HC, was taken to indicate non-shared environmental effects, as such alterations emerge despite the genetic and familial background shared between the HR and LR twins [33,54] (**Case I – non-shared environmental effects**). Differences restricted to HR versus HC, with LR showing no significant differences from either group, were interpreted as risk-expression effects, arising from a combination of shared familial vulnerability (genetic and shared environmental factors common to both twins) and additional individual-specific exposures that selectively affect the HR twin [34,55] (**Case II – risk-expression effects**). When both HR and LR significantly differed from HC, but did not significantly differ from each other, the alteration was considered to reflect a familial or genetic marker, consistent with shared liability and potential endophenotypic properties [34,56,57] (**Case III – familial/genetic marker**), as it appears in both twins regardless of clinical expression of the risk. Finally, differences limited to LR versus HC, with HR showing no significant differences from HC, were interpreted as protective or compensatory mechanisms: LR twin exhibits a deviation from HC, absent in the HR twin, despite their shared genetic background and shared environment.

To validate these results given the limited sample sizes, permutation tests (n=5000) were additionally applied for each comparison and each ROI. For the paired comparison (HR vs LR), a sign-flipping permutation test was used, in which the signs of the within-pair differences were randomly flipped at each iteration and the Wilcoxon signed-rank statistic was recomputed. For the unpaired comparisons (HR vs HC, LR vs HC), a label-permutation test was used, in which group membership was randomly reassigned across all subjects and the Mann–Whitney U statistics was recomputed. In all cases, two-tailed permutation p-values were obtained as the proportion of permuted test statistics at least as extreme as the observed one. The effect size associated with each non-parametric comparison was calculated as the rank-biserial correlation, which quantifies the degree of separation between the two ranked distributions. Values were interpreted as negligible (|r| < 0.1), small (∼0.3), medium (∼0.5), or large (≥ 0.7).

## 3. Results

### 3.1 Sample characterization

The HC sample comprised 31 same-sex twin pairs (13 MZ, 18 DZ-SS) aged 9–31 years (**see Table S1**). Due to missing data across modalities, the analysed sample size varied between analyses: all the 31 pairs had complete linguistic features, 29 pairs also had psychological and sMRI data (**see Table S2**), and 27 pairs had complete data across all modalities, including rs-fMRI. This subsample (N = 27 pairs: 11 MZ, 16 DZ-SS) represents the primary reference group, which has been used for both multimodal neuroimaging analyses and for comparisons with the risk-discordant sample. The risk-discordant sample comprised 17 pairs (9 MZ, age range 10–27 years; 8 DZ-SS, age range 10–24 years) with complete multimodal data. **Table 1** reports the demographic and behavioural characterization of the HC subsample with complete multimodal data (N = 27 pairs) and the risk-discordant group (N = 17 pairs).

### 3.2 Genetic and Environmental Influences on features

In the following sections, the results concerning environmental and genetic influences on psychological, linguistic, and multimodal MRI features are presented. Statistical twin analyses were conducted separately for each feature, considering for each feature set the largest available HC subsample with complete data for that set (n=29 for psychological, n=31 pairs for linguistic, n=27 pairs for neuroimaging features).

#### Psychological features

Among the 17 scales examined in the 29 HC twin pairs, seven met the applicability criterion for analysing phenotypic sources of variance (rMZ>rDZ>0). These included MIS, HPS, ABCL/CBCL Total, ABCL/CBCL Internalizing, ABCL/CBCL subcales related to Somatic Complaints, Thought Problems, Rule-Breaking (**Table 2**. Details on the applicability of the variance decomposition criteria across all psychological scales are reported in **Table S3**.

Five scales showed an ACE variance decomposition. The ABCL/CBCL total score showed the largest heritability (*A* = 0.59) with smaller contributions from shared (*C* = 0.17) and unique (*E* = 0.24) environmental factors. MIS showed balanced genetic and environmental influences (*A* = 0.44, *C* = 0.20, *E* = 0.36), as did the ABCL Rule-Breaking Behaviour subscale, which showed moderate additive genetic (A = 0.49), shared environmental (C = 0.22), and unique environmental (E = 0.29) contributions. In contrast, HPS (*A* = 0.09, *C* = 0.64, *E* = 0.26) and ABCL/CBCL Thought Problems (*A* = 0.15, *C* = 0.66, *E* = 0.18) were predominantly influenced by environmental factors, particularly the shared environment.

Two scales showed an ADE variance decomposition. ABCL/CBCL Internalizing Problems showed broad heritability (*A + D* = 0.52), with contributions from additive genetic (*A* = 0.23) and dominant genetic (*D* = 0.29) effects, alongside substantial unique environmental influence (*E* = 0.48). Similarly, ABCL/CBCL subscales of Somatic Complaints showed a broad heritability (*A + D* = 0.62), comprising additive (*A* = 0.26) and dominant (*D* = 0.36) genetic effects, with the remaining variance attributable to unique environmental factors (*E* = 0.38).

#### Linguistic features

Among the nine linguistic features examined in the 31 HC twin pairs, only narrative fluency satisfied both applicability conditions (rMZ>rDZ>0), showing an epistatic effect (rMZ > 4rDZ; rMZ = 0.82, rDZ = 0.20) (**Table 2**). Details on linguistic features not decomposed into genetic and environmental components are reported in **Table S4**.

#### sMRI features

Among the 8 GMV measures examined in the 29 HC twin pairs, 5 satisfied both applicability conditions (rMZ>rDZ>0). Of these, 4 were best explained by ACE decomposition (the right MFG, the right triangular IFG, and bilateral STG) and 1 showed an epistatic pattern — the left opercular IFG **(Table 3**). The applicability status of the remaining GMV measures is summarized in **Table S5**.

Among the regions influences by ACE sources of variances, the right MFG showed a predominant environmental contribution (C + E = 0.90), mainly driven by unique environmental factors (E = 0.50; C = 0.40), with a lower heritability (A = 0.10). Similarly, the right triangular IFG was primarily influenced by environmental factors (C + E = 0.81), with shared environmental influences accounting for most of the variance (C = 0.51, E = 0.30; A = 0.19). The right STG also showed a strong environmental contribution (C + E = 0.77), largely attributable to shared environmental factors (C = 0.57, E = 0.20), whereas the left STG displayed a more balanced profile, with additive genetic effects accounting for 47% of the variance (A = 0.47) and environmental factors accounting for the remaining 53% (C = 0.27, E = 0.26).

The left opercular IFG exhibited an epistatic pattern, characterized by a markedly stronger correlation in MZ than DZ twins (rMZ = 0.68, rDZ = 0.09), suggesting the involvement of non-additive genetic mechanisms.

Among the 8 CT measures examined in 29 HC twin pairs, only 2 satisfied both applicability conditions (rMZ > rDZ > 0). The left STG was best explained by ACE decomposition, showing a predominant environmental contribution (C + E = 0.69), primarily driven by shared environmental factors (C = 0.46), alongside lower heritability (A = 0.31) and unique environmental effects (E = 0.23). The left pars opercularis of the IFG was best explained by ADE decomposition, with a broad heritability accounting for more than half of the variance (A + D = 0.61), largely driven by dominant genetic factors (D = 0.53), with modest additive genetic effects (A = 0.08) and unique environmental influences accounting for the remaining variance (E = 0.39) (**Table 3**). The corresponding applicability information for the remaining CT measures is reported in **Table S5**.

#### fMRI features

Among the fMRI-derived EC features extracted in 27 HC pairs, only the node-level Netflow of the right triangular IFG satisfied both applicability conditions (rMZ > rDZ > 0). This feature was best explained by an ACE decomposition, with a predominant unique environmental contribution (E = 0.65), negligible shared environmental influence (C = 0.01), and lower heritability (A = 0.35) (**Table 3**). Additional details on the applicability of the variance decomposition criteria for node-level NetFlow features are reported in **Table S6**.

### 3.3 Association of multimodal language-related brain phenotypes with psychopathological risk and zygosity

#### sMRI features

For GMV, in the within-pair analysis, no significant associations emerged between intra-pair differences and psychopathological discordance across any ROI. However, a significant negative correlation was observed between intra-pair GMV differences in the left STG and zygosity (partial r = −0.32, p = 0.03), when controlling for discordance status. This suggests that genetic factors contribute to preserving structural similarity in the left STG, as MZ twins showed greater intra-pair GMV homogeneity than DZ twins independently of psychopathological risk status (**Figure 1B**).

**Figure 1.**
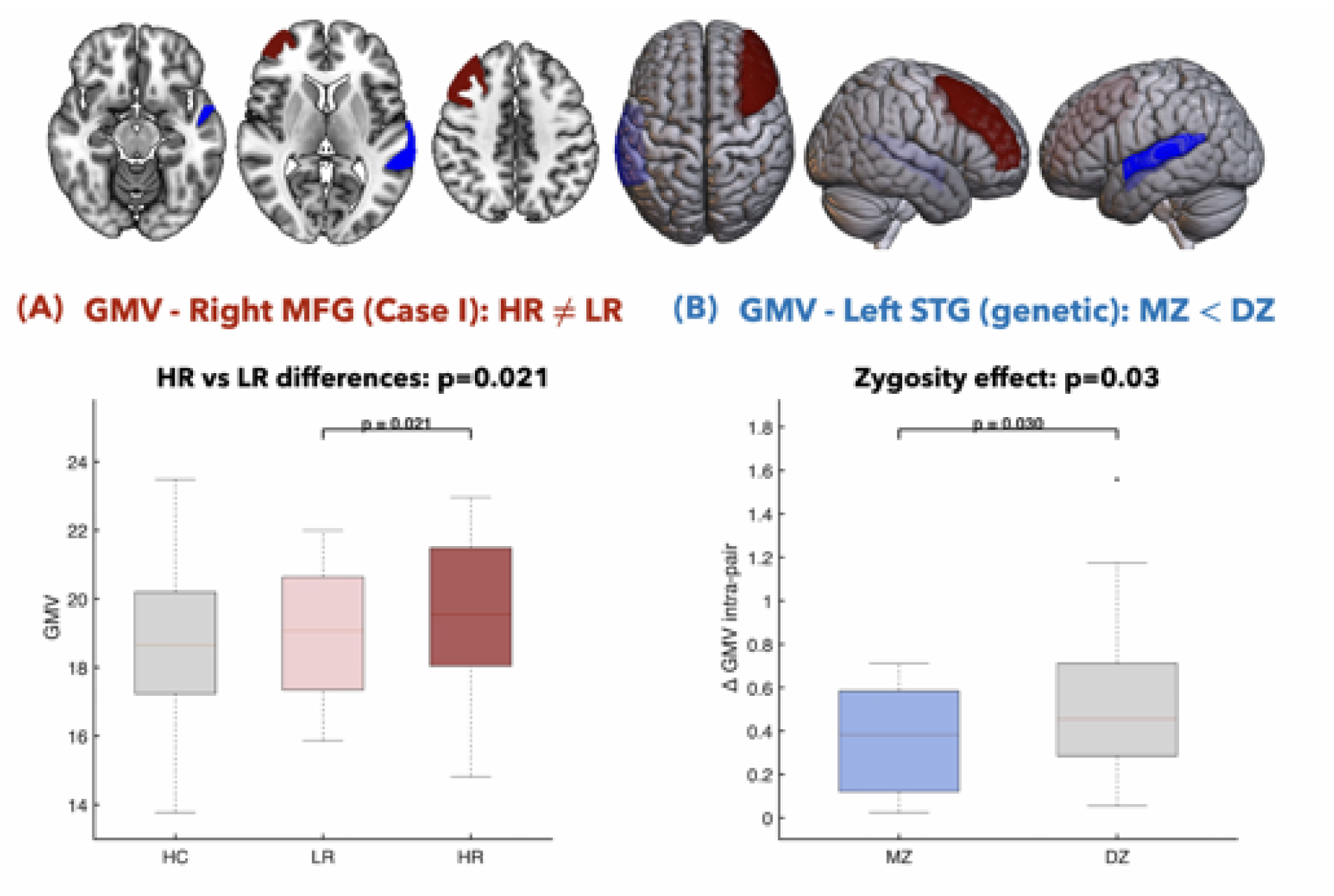
Significant GMV findings in risk-discordant and healthy twin pairs. **(A)** Right middle frontal gyrus (MFG, red): across-group comparison showing significantly increased gray matter volume (GMV) in high-risk (HR) twins relative to their low-risk (LR) co-twins (Case I: HR ≠ LR; Wilcoxon signed-rank test: p = 0.021, p-permuted = 0.022; rank-biserial correlation *effect size* = 0.63). **(B)** Left superior temporal gyrus (STG, blue): within-pair analysis showing significantly greater intra-pair GMV differences in dizygotic (DZ) compared to monozygotic (MZ) twin pairs (partial r = −0.32, p = 0.03, controlling for discordance status), suggesting a genetic contribution to structural homogeneity in this region independent of psychopathological risk discordance. Top panels display the anatomical localization of each region; bottom panels show boxplots of individual GMV values across HC, LR, and HR groups (A) and absolute intra-pair GMV differences across MZ and DZ pairs (B). GMV: gray matter volume; MFG: middle frontal gyrus; STG: superior temporal gyrus; HR: high-risk twin; LR: low-risk twin; HC: healthy control; MZ: monozygotic; DZ: dizygotic.

In the across-group comparison among HR, LR, and HC, only the right MFG showed a statistically significant difference (Table 4): HR twins presented increased GMV relative to their LR co-twins (Wilcoxon signed-rank test: p = 0.021, p-permuted = 0.022; rank-biserial correlation effect size = 0.63, large effect; Table 4), consistent with Case I (non-shared environmental effects; **Figure 1A**).

**Table 4.** Group-level descriptive statistics and pairwise comparisons of GMV across HR twins, their LR co-twins, and HC groups for the eight language-related ROIs (risk-discordant pairs: N = 17 twin pairs; HC: N = 27 twin pairs).

|  | Risk-discordant MZ and DZ twin pairs |  |  |  | Healthy control twin pairs |  |  |
| --- | --- | --- | --- | --- | --- | --- | --- |
| ROI | HR (n=17) |  | LR (n=17) |  | HC (n=27) |  | Significant comparisons |
|  | Mean ± SD | Median | Mean ± SD | Median | Mean ± SD | Median |  |
| Left MFG | 19.43 ± 2.56 | 19.20 | 19.02 ± 2.21 | 19.27 | 18.34 ± 2.45 | 18.51 | *n.s.* |
| Right MFG | 19.46 ± 2.42 | 19.54 | 18.83 ± 2.02 | 19.06 | 18.73 ± 2.51 | 18.66 | HR vs LR: p=0.021, p_perm=0.022, effect size=0.63 |
| Left opercular IFG | 3.34 ± 0.45 | 3.35 | 3.29 ± 0.35 | 3.44 | 3.19 ± 0.45 | 3.13 | *n.s.* |
| Right opercular IFG | 3.48 ± 0.45 | 3.47 | 3.38 ± 0.31 | 3.49 | 3.39 ± 0.50 | 3.42 | *n.s.* |
| Left triangular IFG | 3.57 ± 0.46 | 3.53 | 3.50 ± 0.29 | 3.53 | 3.36 ± 0.49 | 3.28 | *n.s.* |
| Right triangular IFG | 3.59 ± 0.44 | 3.59 | 3.43 ± 0.32 | 3.44 | 3.48 ± 0.53 | 3.51 | *n.s.* |
| Left STG | 6.91 ± 0.76 | 6.96 | 6.77 ± 0.64 | 6.91 | 6.77 ± 0.91 | 6.97 | *n.s.* |
| ROI | HR (n=17) |  | LR (n=17) |  | HC (n=27) |  | Significant comparisons |
| | Mean ± SD | Median | Mean ± SD | Median | Mean ± SD | Median | |
| Right STG | 7.21 ± 0.78 | 7.08 | 7.14 ± 0.68 | 7.12 | 7.03 ± 0.94 | 7.09 | *n.s.* |

Values are reported as mean ± standard deviation and median for the eight language-related ROIs. HR vs LR comparisons were performed using the Wilcoxon signed-rank test (paired); HR vs HC and LR vs HC comparisons were performed using the Mann–Whitney U test (unpaired). Abbreviations: ROI, region of interest; *p*<sub>perm</sub>: permutation-based *p*-value (5000 permutations); *n.s.*: not significant. GMV: gray matter volume; HR: high-risk twin; LR: low-risk twin; HC: healthy control; MFG: Middle Frontal Gyrus; IFG: inferior frontal gyrus; STG: Superior Temporal Gyrus; *n.s.*, not significant.

Since HR and LR twins are co-twins sharing the same familial background, this difference may reflect individual-specific environmental exposures selectively affecting the HR twin, though the inclusion of both MZ and DZ pairs should be noted as a limitation of this interpretation. No other statistically significant group differences were observed across the remaining ROIs (**Table 4**).

Visualization of intra-pair GMV differences across all eight ROIs is provided in **Figures S1–S2**.

For CT, the within-pair analysis revealed no significant associations with either discordance or zygosity. In the across-group comparison, a nominally significant CT increase was observed in the right MFG in HR relative to HC (Wilcoxon rank-sum test: p = 0.04, p-permuted = 0.05; rank-biserial correlation effect size = 0.35, medium effect; **Table 5**), consistent with Case II (risk-expression effects), suggesting a vulnerability shaped by both genetic predisposition and individual-specific environmental factors. In **Figure 2**, we reported the individual CT values of the right MFG across HR, LR, and HC groups. Visualization of intra-pair CT differences across all eight ROIs is provided in **Figures S3–S4**.

**Figure 2.**
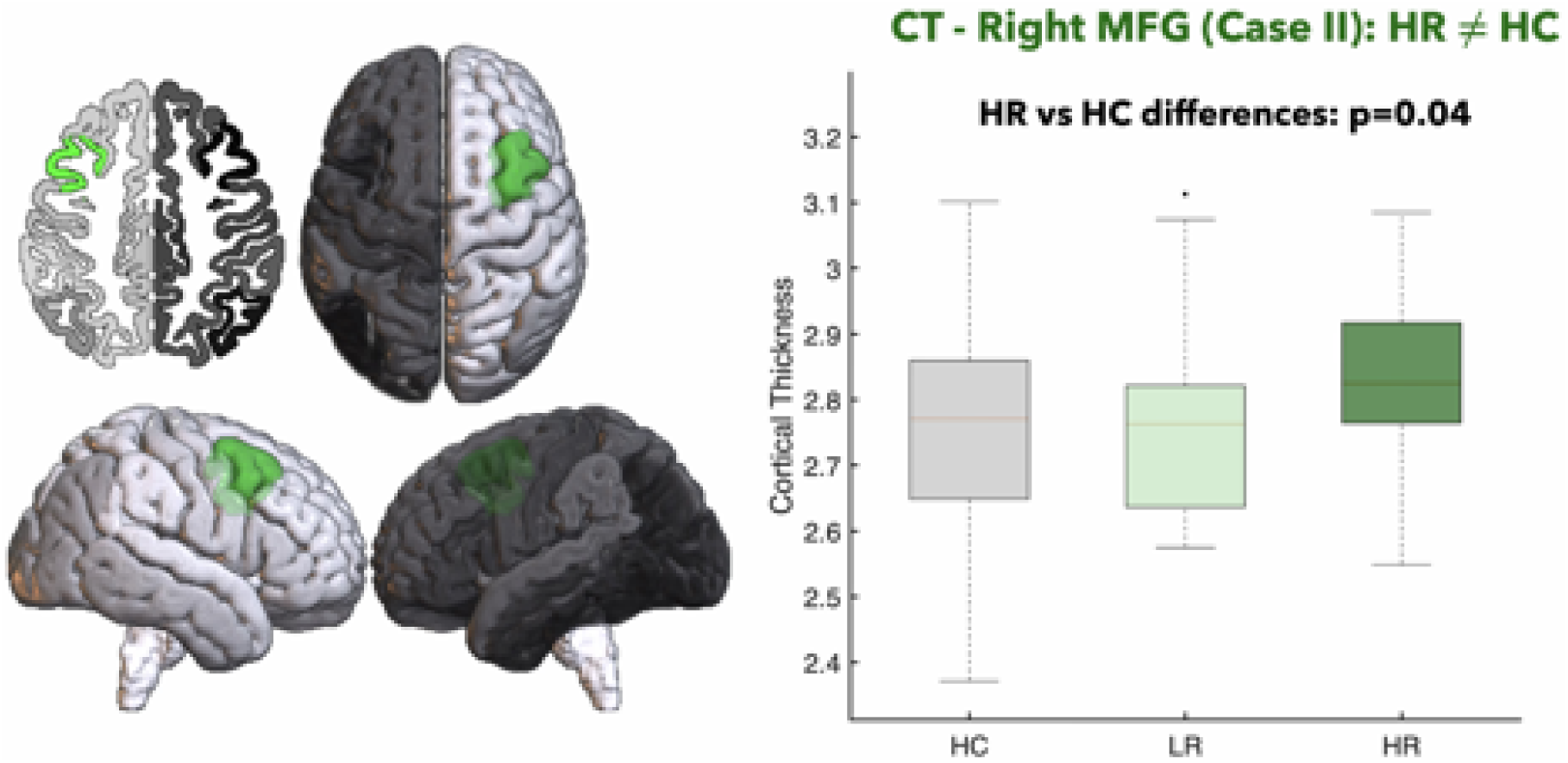
Significant CT finding in risk-discordant and healthy twin pairs. The right middle frontal gyrus (MFG, green) showed increased cortical thickness (CT) in high-risk (HR) individuals relative to healthy controls (HC) in the across-group comparison (Case II: HR ≠ HC; Wilcoxon rank-sum test: p = 0.04, p-permuted = 0.05; rank-biserial correlation effect size= 0.35), with low-risk (LR) twins not differing significantly from either group. Left panels display the anatomical localization of the right MFG; right panel shows individual CT values across HC, LR, and HR groups. CT: cortical thickness; MFG: middle frontal gyrus; HR: high-risk twin; LR: low-risk twin; HC: healthy control.

**Table 5.** Group-level descriptive statistics and pairwise comparisons of CT across HR twins, their LR co-twins, and HC groups for the eight language-related ROIs (risk-discordant pairs: N = 17 twin pairs; HC: N = 27 twin pairs).

|  | Risk-discordant MZ and DZ twin pairs |  |  |  | Healthy control twin pairs |  |  |
| --- | --- | --- | --- | --- | --- | --- | --- |
| ROI | HR (n=17) |  | LR (n=17) |  | HC (n=27) |  | Significant comparisons |
| | Mean ± SD | Median | Mean ± SD | Median | Mean ± SD | Median | |
| Left MFG | 2.827 ± 0.160 | 2.909 | 2.815 ± 0.183 | 2.751 | 2.763 ± 0.157 | 2.766 | *n.s.* |
| Right MFG | 2.825 ± 0.135 | 2.825 | 2.776 ± 0.163 | 2.763 | 2.743 ± 0.151 | 2.771 | HR vs HC: *p*=0.04, *p*<sub>perm</sub>=0.05, effect-size=0.35 |
| Left opercular IFG | 2.857 ± 0.171 | 2.831 | 2.851 ± 0.195 | 2.881 | 2.851 ± 0.178 | 2.887 | *n.s.* |
| Right opercular IFG | 2.822 ± 0.165 | 2.830 | 2.780 ± 0.181 | 2.811 | 2.814 ± 0.113 | 2.811 | *n.s.* |
| Left triangular IFG | 2.786 ± 0.157 | 2.784 | 2.787 ± 0.226 | 2.778 | 2.728 ± 0.147 | 2.712 | *n.s.* |
| Right triangular IFG | 2.779 ± 0.155 | 2.770 | 2.830 ± 0.166 | 2.842 | 2.796 ± 0.155 | 2.778 | *n.s.* |
| Left STG | 2.849 ± 0.161 | 2.855 | 2.849 ± 0.214 | 2.835 | 2.843 ± 0.171 | 2.834 | *n.s.* |
| Right STG | 2.958 ± 0.166 | 2.970 | 2.905 ± 0.219 | 2.860 | 2.960 ± 0.189 | 2.990 | *n.s.* |

Values are reported as mean ± standard deviation and median for the eight language-related ROIs. HR vs LR comparisons were performed using the Wilcoxon signed-rank test (paired); HR vs HC and LR vs HC comparisons were performed using the Mann–Whitney U test (unpaired). Abbreviations: ROI, region of interest; *p*<sub>perm</sub>: permutation-based *p*-value (5000 permutations); *n.s.*: not significant. CT: cortical thickness; HR: high-risk twin; LR: low-risk twin; HC: healthy control; MFG: Middle Frontal Gyrus; IFG: Inferior Frontal Gyrus; STG: Superior Temporal Gyrus; *n.s.*, not significant.

#### fMRI features

For fMRI-derived EC features, neither the within-pair nor the group-level analysis revealed significant differences across any ROI (**Table 6**). Visualizations of intra-pair differences and across-group profiles across all eight ROIs are provided in **Figures S5–S6**.

**Table 6.**
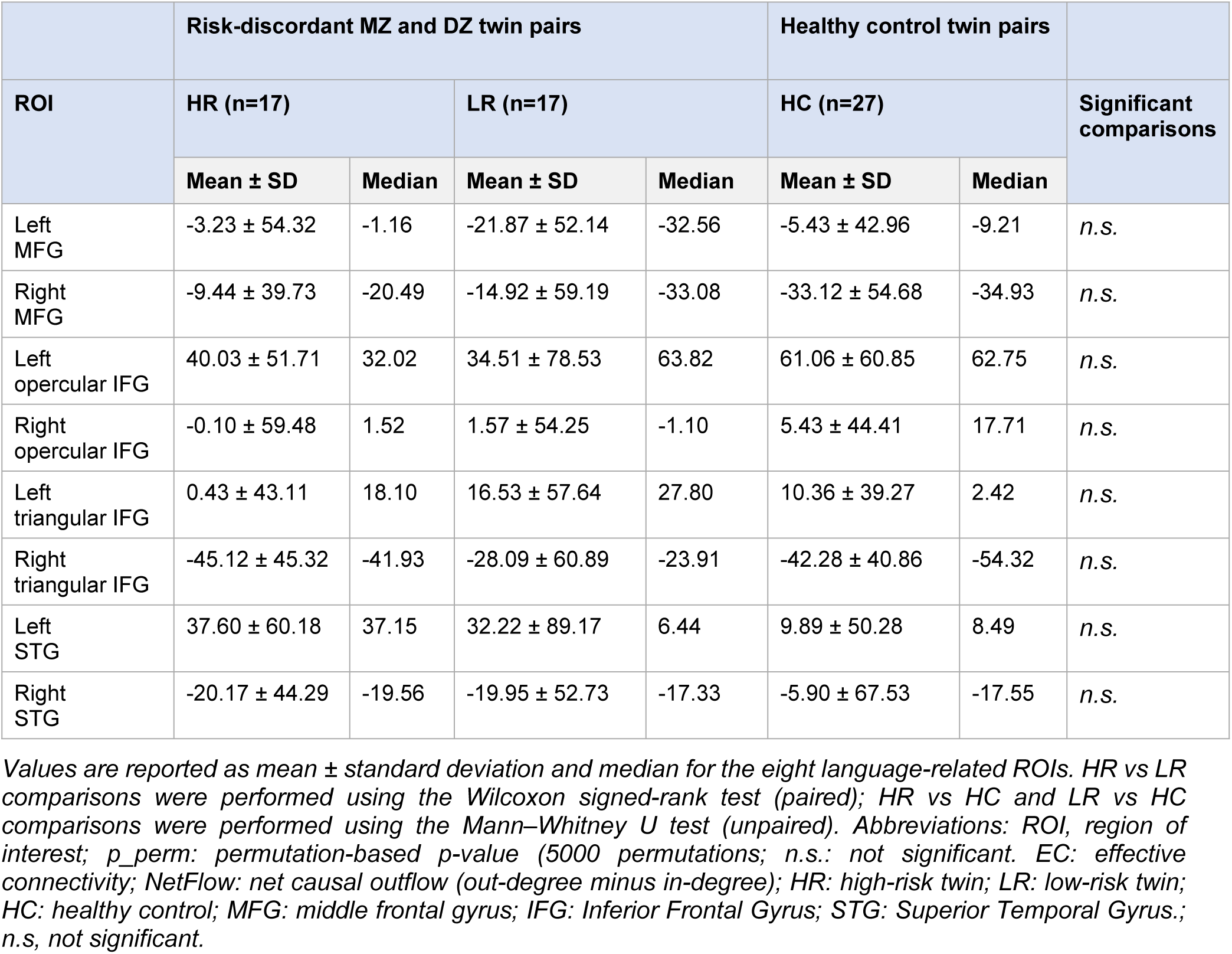
Group-level descriptive statistics and pairwise comparisons of NetFlow across HR twins, their LR co-twins, and HC groups for the eight language-related ROIs (risk-discordant pairs: N = 17 twin pairs; HC: N = 27 twin pairs).

## 4. Discussion

In our study, a cross-sectional multimodal dataset of young pairs of twins was analysed to examine how genetic and environmental determinants shape psychological, linguistic, and brain features in language-related brain regions during adolescence and young adulthood.

Multimodal brain phenotypes were analysed within language-related brain regions and included structural features of GMV, CT, and node-level EC as functional patterns. For the first time, genetic and environmental determinants were comprehensively explored by integrating univariate twin statistical analyses in healthy twins with within-pair and group-level comparisons in twin pairs discordant for psychopathological risk, across structural and functional neuroimaging measures alongside linguistic and emotional-behavioural domains. By combining these analytical approaches, we investigate whether GMV, CT, and functional EC measures within language.related brain regions are shaped by nature and nurture and how structural and functional alterations in these regions are associated with psychopathological risk.

As a first step, univariate twin statistical analyses in healthy twins characterized the balance between genetic and environmental influences across psychological measures, linguistic features, and multimodal neuroimaging markers in eight language-related brain regions. These analyses revealed that, in the absence of psychopathological risk, variability in these regions are largely driven by environmental factors across both structural and functional brain measures, with only limited and region-specific genetic contributions. Notably, for cortical structure and functional patterns, the evidence is regionally specific rather than global: the heritability of CT varies substantially by region [19], and twin imaging studies show that cortical thickness and white-matter connectivity can share genetic influences in selected networks[21]. These observations underscore the importance of interpreting genetic and environmental effects on a region-by-region basis rather than assuming uniform environmental influences across all language-related cortex. Regarding psychological phenotypes, shared environmental factors showed particularly strong influences on thought problems, while rule-breaking behaviour showed a more balanced genetic and environmental profile. Hypomanic personality traits also showed a predominant shared environmental contribution, while magical ideation showed moderate genetic influences alongside environmental contributions, further supporting the relevance of environmental factors across multiple phenotypic domains beyond neuroimaging.

In contrast, narrative fluency [4,40] showed strong epistatic genetic effects, consistent with evidence of heritability for higher-order language abilities, consistent with evidence of heritability for higher-order language abilities[27]. Moderate-to-high heritability was also observed for selected pscyhological measures, specifically ABCL/CBCL internalizing symptoms, somatic complaints, and the total score under ADE and ACE variance decomposition, consistent with previous CBCL-based twin studies reporting moderate-to-high heritability across emotional-behavioural dimensions [29–31].

In addition to this univariate statistical twin analyses, in twin pairs discordant for psychopathological risk we tested whether and how alterations in multimodal brain features are associated with the considered risk. This approach allowed distinguishing baseline neurobiological variability from alterations emerging in association with risk expression. Consistent with the univariate framework, genetic effects contributed only to a small subset of regions and imaging modalities, whereas risk-related structural alterations appear to reflect either individual-specific environmental influences or their interaction with shared familial liability. The right MFG carries the clearest GMV and CT alterations, while the left STG appears more stable and may reflect familial or genetic effects on structural homogeneity rather than direct risk expression. This evidence is consistent with twin MRI studies in schizophrenia showing that some GMV abnormalities track illness or unique environmental exposure beyond genetic liability alone [33,34]. Converging evidence from affective-disorder twin studies indicates that both genetic and environmental factors shape brain vulnerability through distinct mechanisms, including environmentally driven amygdala hyperreactivity in monozygotic discordant pairs[58], gene-environment influences on amygdalar resting-state connectivity [59], genetic liability associated with regional gray matter reductions [60].

By integrating findings from within-pair and group-level analyses across GMV, CT, and functional EC patterns, the present study suggests that early neurobiological vulnerability in language-related brain regions emerge primarily from environmental effects rather than genetic factors. If confirmed on larger samples, these results could inform on the balance between genetic and environmental effects on multimodal brain features within language-related networks during a critical developmental period. This knowledge might ultimately prove useful in understanding susceptibility factors for mental illnesses that arise during adolescence and young adulthood, with specific relevance to brain regions supporting language and higher-order cognitive functions.

### 4.1 Genetic and Environmental influences on features

Univariate twin analyses conducted in HC twin pairs showed that linguistic, psychological, and neurobiological traits are differentially influenced by genetic and environmental factors. While linguistic and psychological features showed moderate to strong genetic influences, multimodal neuroimaging measures within language-related regions were predominantly shaped by shared and unique environmental factors, a pattern especially evident for GMV and EC. Overall, these results indicate that structural and functional brain variability in language-related regions is weakly genetically determined and highly sensitive to environmental influences.

#### 4.1.1 Genetics

Our findings suggest limited and highly selective genetic influences on neuroimaging features, alongside stronger genetic contributions to specific linguistic and psychological traits, pointing to different nature-nurture contributions between genetic architecture of behavioral phenotypes and that of brain structure and function in language-related brain regions.

Among neuroanatomical measures, only the CT of the left pars opercularis of the IFG — corresponding to Brodmann area 44, a core component of Broca’s area [61] — was best explained by an ADE variance decomposition, indicating both additive and non-additive genetic contributions. Similarly, GMV of the left opercular IFG resulted to be influenced by epistatic effect (rMZ = 0.68, rDZ = 0.09), suggesting the involvement of non-additive genetic mechanisms. The convergence of genetic effects across both CT and GMV in this region points to the left opercular IFG as a region of particular genetic interest within the language brain network, where both structural volume and thickness appear influenced by non-additive genetic factors. In contrast, all remaining GMV features meeting the applicability criteria were predominantly explained by ACE variance decomposition with dominant environmental contributions, and no genetic effects were detected for directed functional patterns derived from EC.

Regionally specific genetic influences on CT have been well documented[19], and some twin studies have shown that CT and associated white-matter connectivity share genetic influences in selected networks[21], further supporting a region-by-region rather than global interpretation of genetic effects.

Interestingly, the review by van der Meer & Kaufmann (2022)[20] highlighted substantial heterogeneity in genetic expression across cortical regions and developmental stages, which suggests that structural measures may be particularly susceptible to environmental modulation during the young adulthood period. While our results mainly point to an overall larger environmental effect on structural measures in language-related regions, some studies have shown stronger genetic contributions. Specifically, Yoon et al. (2010)[62] reported strong genetic influences on CT in the MFG in a homogeneous sample of 8-year-old twins. This inconsistency may be partially explained by the wider age range of our sample, as previous research has shown that the influence of genetic and environmental factors on brain structure changes across development [17,24].

At the linguistic and psychological level, genetic influences were more prominent. Although previous studies have demonstrated moderate heritability for higher-order language abilities[27] and genetic overlap with reading comprehension[25], productive language measures are typically dominated by nonshared environmental influences [26]. The epistatic genetic effect observed for narrative fluency suggests that the overall efficiency and structure of discourse production may carry a stronger heritable component than other productive language skills, a finding that warrants replication in larger samples. Among the psychological measures derived from the ABCL/CBCL, internalizing symptoms and somatic complaints showed broad heritability under ADE decomposition (H² = 0.52 and H² = 0.62, respectively), while the total score showed moderate narrow heritability under ACE decomposition (h² = 0.60). These estimates are broadly consistent with previous twin studies in children and adolescents focusing on emotional-behavioural domain, which have reported moderate-to-high heritability across emotional-behavioural dimensions, with estimates typically ranging from 0.50 to 0.70 for internalizing, externalizing, and DSM-oriented scales[29–31].

#### 4.1.2 Environment

Environmental factors emerged as the predominant source of variability across multimodal neuroimaging features. All GMV measures meeting applicability criteria were best explained by ACE phenotypic variance with larger environmental components, with shared environmental effects especially prominent for the right triangular IFG and right STG, and substantial unique environmental contribution for the right MFG. These findings are consistent with Alijanpourotaghsara et al. (2025) [22], who reported that in healthy adult twins, frontal and temporal regions are mostly influenced by environmental factors, while genetic effects predominate in parietal and occipital regions.

Environmental influences were also evident for CT in the left STG, with a predominance of shared environmental effects, supporting the sensitivity of temporal language-related cortices - including Wernicke’s area - to environmental input during development [63]. This is broadly consistent with longitudinal twin evidence showing shared environmental contributions to cortical development in superior temporal regions during childhood and adolescence, where CT showed lower heritability relative to surface area[23].

At the functional level, node-level measures of EC showed a strong environmental signature, with the functional measure of NetFlow right triangular IFG predominantly influenced by unique environmental factors. The right IFG is involved in prosodic and paralinguistic aspects of language [13] and may play a greater role in early development, when language networks are less strongly left-lateralized [64], suggesting that the observed environmental influence may reflect experience-dependent reorganization of functional networks. Of note, these findings extend those from our previous twin study on the same cohort[18], which showed a predominant role of unique environment on multi-scale resting-state functional connectivity. Although in this study we consider as functional brain features the directed EC measures focused on language-specific brain circuits, we replicate the environmental predominance providing complementary evidence across two distinct functional neuroimaging approaches.

Among emotional-behavioural psychological measures, shared environment showed a particularly strong influence on thought problems, while rule-breaking behaviour showed a more balanced profile, with moderate genetic and environmental contributions. This is consistent with previous twin studies reporting substantial environmental contributions to specific CBCL dimensions, including anxiety/depression and delinquent behaviour, although the relative balance between genetic and environmental effects varies by sex, age, and behavioural phenotype [29,30]. Of note, the HPS and MIS, measures of subclinical psychosis-proneness traits, also showed substantial environmental contributions alongside moderate genetic influences. These findings are broadly consistent with previous twin studies reporting predominant environmental effects on both hypomanic personality and magical ideation dimensions, with genetic contributions varying substantially across samples, sex, and developmental stage[16,65]. To our knowledge, direct twin estimates in adolescent samples remain limited, and these preliminary findings warrant replication in larger independent cohorts.

Overall, this pattern highlights the importance of both shared and individual-specific environmental exposures in modulating brain organization and behaviour in healthy adolescents and young adults. Structural and functional brain variability in language-related regions remains highly sensitive to environmental exposures, while among emotional-behavioural psychological traits environmental effects are selectively prominent for thought problems and hypomanic personality. Together with previous longitudinal twin evidence showing stable genetic effects on internalizing and externalizing trajectories across development [32], our findings suggest that emotional-behavioural traits arise from varying combinations of genetic liability and environmental exposure, with the relative contribution depending on the specific behavioural domain.

### 4.2 Multimodal language-related brain phenotypes associated with psychopathological risk and zygosity

Analyses in risk-discordant twin pairs provide convergent evidence that non-shared environmental influences represent the primary driver of neuroanatomical variability associated with psychopathological risk, while genetic effects appear limited and region-specific. The significant findings converge on two regions: the right MFG, where both GMV (Case I: HR vs LR) and CT (Case II: HR vs HC) showed alterations consistent with non-shared environmental effects and risk expression, and the left STG, which was the only region where a significant association between intra-pair GMV differences and zygosity pointed to a genetic contribution, suggesting a stabilizing role on structural homogeneity rather than a link to psychopathological vulnerability. No significant resting-state directed connectivity effects were observed in either the within-pair or the across-group analyses; this may partly reflect the resting-state nature of the fMRI data, in which directed connectivity measures may be less sensitive to risk-related modulations in the absence of a language task[66]. However, the limited sample size of discordant pairs constrained statistical power, and results should be interpreted accordingly.

#### 4.2.1 Genetic effects

In the within-pair analysis, intra-pair GMV differences in the left STG were significantly associated with zygosity but not with psychopathological discordance. This suggests that genetic factors may contribute to preserving structural similarity in the left STG across twin pairs, independently of psychopathological risk status. Of note, previous studies have reported that the left STG, specifically in the planum temporal and Heschl gyrus, both in first episode schizophrenia[67] and with progressive gray matter loss over time[68], suggesting that the morphology of this region may carry a familial or biological vulnerability. Our finding of a zygosity-related effect on GMV homogeneity is consistent with this view, pointing to a genetic contribution to structural stability in this region independent of psychopathological risk expression. Importantly, no comparable genetic effects were observed for any other GMV region, or for CT or EC measures.

#### 4.2.2 Unique environmental effects

The statistically significant finding points to a role of non-shared environmental influences in shaping GMV differences associated with psychopathological risk.

The clearest evidence emerged from the comparison of individual neuroimaging values across HR, LR, and HC groups for GMV in the right MFG, where HR twins showed significantly increased volume compared to their LR co-twins (Case I: HR vs LR). Since this difference appears between co-twins sharing the same familial background, it points to individual-specific environmental exposures as the likely source of this alteration, though this interpretation should be considered cautiously as the sample included both MZ and DZ pairs, and therefore does not fully isolate non-shared environmental effects from genetic differences between DZ co-twins. A comparable pattern has been reported by Borgwardt et al. (2010)[33] in twins discordant for schizophrenia: affected twins showed reduced GMV in frontal cortical regions compared to their unaffected co-twins, while unaffected co-twins did not differ from healthy controls. The authors concluded that these alterations were not attributable to genetic liability alone but might also reflect environmental factors. Notably, Cannon et al. (2002)[34] used cortical surface mapping in twins discordant for schizophrenia and identified disease-specific gray-matter deficits primarily in dorsolateral prefrontal cortex, superior temporal gyrus, and superior parietal lobule, while genetic proximity to an affected twin was associated with deficits primarily in prefrontal cortex — demonstrating that genetic and disease-specific influences affect gray matter in partially non-overlapping areas. If confirmed in larger samples, this evidence may inform on the environmental susceptibility factors underlying neurobiological vulnerability during adolescence.

#### 4.2.3 Familial liability and risk expression

A second class of findings appears to reflect risk expression arising from the interaction between shared familial liability and individual-specific environmental exposures. This pattern was most evident in right frontal regions, where structural alterations reached significance.

In the comparison of individual neuroimaging values across HR, LR, and HC groups, CT in the right MFG showed a significant increase in HR individuals relative to HC, with LR twins not differing from either group (Case II: HR vs HC). Unlike Case I, this pattern does not show a clear HR–LR separation and therefore cannot be interpreted as an effect created primarly from non-shared environmental effect. This pattern suggests that this cortical alteration is more closely related to risk expression, arising from a combination of shared familial vulnerability and additional individual-specific exposures that selectively affect the HR twin. Notably, the direction of this alteration, increased rather than decreased CT in HR individuals, is consistent with evidence linking perceived stress to increased rostral MFG cortical thickness in family-based and discordant-sibling designs [69], suggesting that individual-specific environmental exposures may contribute to cortical thickening in this prefrontal region.

In this context, Ettinger et al. (2012)[70] studied MFG volume in monozygotic twins concordant and discordant for schizophrenia. They found MFG volume reductions in concordant pairs — where both twins are affected and share maximal familial loading — but not in the affected twin from discordant pairs. The absence of MFG alterations in Ettinger’s discordant affected twins might seem in contrast with our evidence reporting CT alterations in the HR twin of discordant pairs. In this respect, it should be noted that this discrepancy likely reflects differences in the measure examined and in illness stage: Ettinger examined established schizophrenia, whereas our study may capture an earlier stage of risk. This suggests that structural MFG alterations in established illness may require stronger shared familial loading, while at earlier stages of risk, as in our twin sample, individual-specific environmental factors may play a more prominent role. Nonetheless, this interpretation is also supported by fMRI and network-based studies reporting altered functional integration and hypoactivation of the right MFG in individuals at clinical high risk or in early stages of schizophrenia, suggesting that this region may reflect early susceptibility expression rather than a genetically fixed lesion site [71].

Together, both GMV and CT findings in the right MFG capture a role of individual-specific environmental exposures, differing in that the GMV alteration emerges exclusively within pairs, supporting a more prominent individual-specific effect, while the CT alteration, observed in HR relative to HC but not within pairs, additionally points to an interaction with shared familial liability. Both findings converge on the right MFG as a region of heightened neurodevelopmental sensitivity, where genetic predisposition alone appears insufficient to account for the observed alterations.

#### 4.2.4 Study limitations

The present study has some limitations that need to be acknowledged. The major limitation is the small sample size, which limits statistical power and constrains the interpretation of results. As in our previous study [18], the limited number of twin pairs prevented us from fitting Structural Equation Models (SEM), which would have offered more robust hypothesis testing, including confounding and moderation effects on genetic and environmental estimates, as well as accounting for measurement error. Instead, heritability estimates were derived from Falconer’s formula, which, although providing valid estimates of the proportion of phenotypic variance attributable to genetic factors, does not allow simultaneous estimation of all variance components. Moreover, in small twin samples, non-additive genetic components may be masked by shared environmental factors, potentially altering heritability estimates.

To take full advantage of the discordant twin design, larger samples are needed. The limited number of risk-discordant pairs (N = 17) constrained statistical power for the within-pair and across-group analyses, and prevented us from stratifying by zygosity, which may have reduced the degree of genetic control and limited interpretability. A larger sample would also have allowed us to reliably detect genetic effects of limited magnitude and to test for common environmental effects, for which the classical twin design is generally underpowered.

Despite these constraints, it should be noted that the study drew on multimodal MRI data from discordant twins drawn from the general population. Furthermore, subjects included in this study were not help seekers since at-risk status was identified through standardized screening instruments (ERIraos-CL and CBCL/ABCL cutoffs) via the Italian Twin Registry. This approach enabled the detection of potential brain alterations at a very early, subclinical stage of psychopathological risk. However, it also implies that our at-risk twins may present milder phenotypes than those typically examined in clinical twin studies, limiting the direct comparability of findings. Additionally, the T-score cutoff used to define psychopathological risk on the CBCL/ABCL remains an arbitrary threshold that does not correspond to a clinical diagnosis and may influence risk classification.

Additional methodological considerations should be mentioned. The wide age range of our sample (10–31 years) spans different developmental stages during which the balance between genetic and environmental influences on brain structure is known to shift, potentially introducing heterogeneity in the estimated effects. Among linguistic features, only narrative fluency met the applicability conditions for twin statistical analyses, limiting our ability to draw broader conclusions about the heritability of language-related traits. Future studies with larger samples, and potentially including more comprehensive linguistic batteries, may help clarify the genetic architecture of language phenotypes during development.

Furthermore, the cross-sectional nature of the study did not allow us to observe longitudinal trajectories of the altered brain features over time. In the absence of longitudinal data, it remains unclear whether the observed patterns reflect stable traits or dynamic developmental processes. Future longitudinal twin investigations would be particularly informative for tracking how genetic and environmental influences on language-related brain regions evolve across adolescence and young adulthood.

## Data availability statement

The psychological and MRI datasets supporting the current study have not been deposited in a public repository because of privacy and ethical restrictions but are available from the corresponding author on request. Matlab codes are available at Opens Science Framework web application link: https://osf.io/sfumq/overview?view_only=882c8f89e2f34d76886267b5fad1bc27.

## Acknowledgments

This work was supported in part by the LearningTwin grant from Italian Ministry of Health (SG-2021-12375585). MN and MM were partially supported by Ricerca Corrente (RC) 2026. PB was partially supported by grants from the Italian Ministry of Education and Research - MUR (‘Dipartimenti di Eccellenza’ Programme 2023–27 - Dept. of Pathophysiology and Transplantation, Università degli Studi di Milano), the Italian Ministry of Health (Project “Hub Life Science” - Diagnostica Avanzata, HLS-DA, PNC-E3-2022-23683266– CUP: C43C22001630001 / MI-0117 / Piano Nazionale Complementare Ecosistema Innovativo della Salute; Ricerca Corrente 2026), by the ERANET Neuron JTC 2023 (ERP-2023-23684211 - ResilNet) and Eranet Neuron JTC 2024 (ER-2024-23684536 - BRAWO Project).

## Ethics approval statement

This study protocol received the approval of Ethics Committee related to the LearningTwin study (SG-2021- 12375585).

## Patient consent statement

All participants provided a written consent to the study protocol.

## Author contribution statement

ET: Data curation; Project administration; Conceptualization, Methodology; Software; Writing - original draft; Writing - review & editing. CC: Methodology review; Writing - original draft; Writing - review & editing. AP: Writing - original draft; Writing - review & editing. LD: Methodology; Writing - original draft; Writing - review & editing. CP: Writing - original draft; Writing - reviefw & editing. SB: Writing - original draft; Writing - review & editing. FS: Writing - original draft; Writing - review & editing. GD: Conceptualization; Writing – review & editing. EM: Conceptualization; Methodology review; Supervision; Software review; Writing - review & editing. CB: Data curation; Writing – review & editing. MM: Data curation; Writing – review & editing. CF: Conceptualization; Methodology review; Writing – review & editing. EM: Conceptualization; Methodology review; Writing – review & editing. MN: Conceptualization; Data curation: Writing – review & editing. PB: Conceptualization; Supervision; Writing - review & editing.

